# Dementia Risk and Dynamic Response to Exercise: Methodology for an Acute Exericise Clinical Trial

**DOI:** 10.1101/2020.08.22.20179564

**Authors:** Dreu White, Casey S John, Ashley Kucera, Bryce Truver, Rebecca J Lepping, Phil Lee, Laura Martin, Sandra A Billinger, Jeffrey M Burns, Jill K Morris, Eric D Vidoni

## Abstract

**Background:** Exercise likely has numerous, meaningful benefits for brain and cognition. However, those benefits and their causes remain imprecisely defined, especially in the context of cognitive disorders associated with aging, such as Alzheimer’s disease (AD). If the brain does benefit from exercise it does so primarily through exposure to brief, “acute” exposures to exercise over a lifetime. Methods: The Dementia Risk and Dynamic Response to Exercise (DYNAMIC) clinical trial seeks to characterize the acute exercise response in cerebral perfusion, and circulating neurotrophic factors in older adults with and without the apolipoprotein e4 genotype (*APOE4*), the strongest genetic predictor or sporadic, late onset AD. DYNAMIC will enroll 60 older adults into a single moderate intensity bout of exercise intervention. We will measure pre- and post-exercise cerebral blood flow using arterial spin labeling, and neurotrophic factors. We expect that *APOE4* carriers will have poor CBF regulation, i.e. slower return to baseline perfusion after exercise, and will demonstrate blunted neurotrophic response to exercise, with concentrations of neurotrophic factors positively correlating with CBF regulation. If exercise-induced changes in perfusion and circulating factors can be detected, DYNAMIC will contribute to our understanding of exercise-induced brain change and potential biomarker outcomes of exercise interventions. Results: Preliminary proof-of-concept findings on 7 older adults and 9 younger adults. We have found that this experimental method can capture CBF and neurotrophic response over a time course, and best practices following exercise. Conclusions: This methodology will provide important insight into acute exercise response and potential directions for clinical trial outcomes. ClinicalTrials.gov NCT04009629

## Background

The number of Americans 65 years and older will double in size over the next 40 years.(1) Aging is often associate with increased cognitive decline.(2) Alzheimer’s disease (AD) is of particular concern to the health care system, with an expected two-fold increase in prevalence over the next 30 years, and high direct and indirect costs of care.(3, 4) We must find effective interventions to reduce the burden of cognitive decline and AD on our society.

There is evidence that risk of age-related cognitive decline, including mild cognitive impairment and dementia, can be reduced by health behavior interventions such as exercise.(5–12) Although the literature is not conclusive,(13–16) there is a growing consensus that common healthy behaviors, and especially exercise, support brain health and cognitive function.(17, 18) A number of potential mechanisms may link exercise with brain health. Increased brain volume,(19) regional neurogenesis,(20) circulating neurotrophic factors,(21) and cerebrovascular reserve (i.e. capacity for response to a stimulus challenge) (22) all have been implicated as mediators of exercise benefits for the brain. If the brain does benefit from exercise it does so primarily through brief, “acute” exposures to exercise over a lifetime.(23)

Due to the considerable benefits of aerobic exercise on cardiovascular function, there is interest in precisely defining cerebrovascular adaptations to aerobic exercise and how those adaptations may support cognitive function.(24, 25) We know that sedentary older adults demonstrate decreasing cerebral blood flow (CBF) over time.(26–29) Older athletes who take time off of training quickly experience reductions in CBF,(30) whereas habitual exercise appears to increase CBF.(26, 28, 31) Heart disease and stroke patients improve CBF while completing their cardiac rehab program.(32, 33) Yet, evidence from prior aerobic exercise intervention trials with otherwise healthy older adults have been mixed, some reporting increased CBF,(9) and others reporting no difference.(34)

In general, these studies have used passive “resting” conditions when measuring CBF and blood biomarkers, rather than employing tasks or experimental challenge (e.g. exercise, task-based fMRI, neuropsychological test) meant to approximate the ecological stressors of daily life. Our own work and others’ demonstrates the importance of measuring the response to challenge, especially to exercise.(22, 35–42) For example, using transcranial Doppler (TCD), we recently described the dynamic change in a proxy measure of CBF with onset of aerobic exercise in young and older individuals.(35) Our work demonstrates that older adults have a noticeable blunting of CBF during a dynamic condition like exercise that was less appreciably different from younger adults during a static, resting, measure.

There is evidence that the Apolipoprotein epslion4 (*APOE4*) single nucleotide polymorphism, the strongest genetic risk factor for late-onset, sporadic AD,(43) leads to modified neurovascular coupling, a leaky blood–brain barrier, angiopathy, and disrupted nutrient transport.(44) *APOE* appears key to maintaining cerebrovascular integrity independent β amyloid deposition.(45) *APOE4* carriers also demonstrate altered (46–48) and generally lower resting CBF especially in regions associated with AD-related change.(29, 49–53) Because exercise has such a strong and reliable benefit for the vascular system *APOE4* carriers, who are at greater vascular risk than non-carriers,(12) may preferentially benefit.(54–56)

There are well-documented differences in cerebral blood flow (CBF) and tissue oxygenation based on age or *APOE* genotype,(29, 48, 49, 57) with deleterious consequences for cognition.(29, 58) The exercise stimulus may counter this through cerebral oxygenation and stimulation of neurotrophics, among other mechanisms. Cerebrovascular endothelial cells respond to the shear force stress on the vessel walls by releasing Brain Derived Neurotrophic Factor (BDNF).(59, 60) Peripheral Vascular Endothelia Growth Factor (VEGF) appears to be essential for running-induced neurogenesis and benefits acute exercise performance and brain blood flow in mice.(61, 62) Measurable increases in VEGF are seen after acute exercise.(38,39) But to date, the literature has not connected CBF, neurotrophins, and APOE genotype in exercising humans, despite convergent data pointing to their intimate involvement in cognitive of - decline. Our project will extend the prior work by directly assessing the relationship of these factors in response to an acute exercise challenge.

We set out to quantify the CBF response to exercise which has the potential to be a valuable measure of cerebrovascular health.(22) This manuscript details the clinical trial methodology and proof-of-concept preliminary data of the Dementia Risk and Dynamic Response to Exercise study (DYNAMIC - NCT04009629). The scientific premise underlying this project is that CBF and blood-based biomarkers such as VEGF, BDNF, and Insulin-like Growth Factor 1 (IGF1) are interrelated mechanisms driving chronic aerobic exercise effects on brain health and cognition.(9, 20, 63) Our single visit clinical trial seeks to characterize the relationship of *APOE4* carrier status with CBF and blood-based biomarkers of brain health. We capture dynamic fluctuations in CBF and blood-based biomarkers in a time-sensitive manner before and after an acute bout of moderate intensity aerobic exercise. Our working hypothesis is that individuals at genetic risk for AD have poor CBF regulation and altered neurotrophic response and that this methodological approach will inform future clinical trial biomarker protocols in the future.

A secondary goal of this manuscript is to detail the protocol design and adaptations of a single visit experimental study as a clinical trial. Changes in 2018 to the Federal Policy for the Protection of Human Subjects (‘Common Rule’) has expanded the definition of a clinical trial to include any investigation in which human subjects are prospectively assigned to an intervention to evaluate the effect on a biomedical or behavioral outcome.(64) As a result, most experimental exercise manipulations are now classified as clinical trials even if not traditionally considered an intervention, resulting in increased scrutiny and more strict standards for protocol and reporting. This manuscript also serves to detail clinical trial adaptations for single visit or short experimental studies that have previously fallen outside the aegis of clinical intervention regulation.

## Methods

### Summary of design

DYNAMIC is a single site, non-randomized, prospectively enrolling trial testing *APOE4-*related response differences to a single, 15-minute bout of moderate intensity aerobic exercise. The study plans to enroll 60 older adults (> = 65 years), approximately balanced for E4 carriage, and up to 20 younger individuals to serve as normative cohort. We hypothesize that *APOE4* carriers will have poor CBF regulation, i.e. slower return to baseline perfusion (reduced area under the curve) after exercise, and will demonstrate blunted neurotrophic response to exercise, with concentrations of neurotrophic factors positively correlating with CBF regulation. We will also explore the relationship of the CBF and neurotrophic responses to cognitive performance.

### Outcomes

As a registered clinical trial, we have identified a single primary outcome and several secondary outcomes of interest. Our primary outcome of interest is global CBF area under the curve (AUC) of the cumulative cerebral blood flow before and after our exercise intervention. Secondary outcomes are change in IGF1, change in VEGF, and change in BDNF from baseline to post intervention. Our exploratory cognitive measures and associated cognitive domains are episodic memory (NIH Toolbox Picture Sequence Memory Test 8+), processing speed (Pattern Comparison Test 7+), and attention and executive function (Flanker Inhibitory Control and Attention Test 12+).

### Recruitment and Eligibility

We significantly reduce participant burden by heavily leveraging the infrastructure of the University of Kansas Alzheimer’s Disease Center (KU ADC), an NIH-designated Alzheimer’s disease research center. The KU ADC follows a Clinical Cohort of 400 individuals with annual cognitive evaluations and prior genetic testing.(65) We recruit younger participants from the local area via social networks and fliers. All individuals provide written, institutionally approved written informed consent according to the Declaration of Helsinki guidance either on the day of the visit or in advance through electronic consenting.

Participants have no changes in memory or thinking, or diagnosis of cognitive impairment. Additional inclusion criteria are: 1) Age 18–85 (inclusive); 2) English speaking; 3) corrected hearing or vision; 4) willingness to have genotyping performed if necessary. Exclusion criteria are: 1) health care provider recommended activity restrictions; 2) prior diagnosis of clinically significant cognitive decline judged on Clinical Dementia Rating(66) or Quick Dementia Rating Scale(67) and AD8(68) equivalent of non-impaired, or similar clinical determination in the prior 6 months; 3) anti-coagulant use; 4) high cardiovascular risk without physician clearance for exercise(69); 5) exercise-limiting pain, musculoskeletal, or metabolic condition; 6) MRI contraindications; 7) clinically significant psychiatric illness or other neurological disorders that have the potential to impair cognition (e.g., Parkinson’s disease, stroke defined as a clinical episode with neuroimaging evidence in an appropriate area to explain the symptoms); 8) myocardial infarction or symptomatic coronary artery disease in the prior 2 years.

Secondary enrollment considerations are sex and E4 carriage. Within the older adult Clinical Cohort 95% already have *APOE* genotype determined from prior study activities and have consented to share that information with collaborating studies such as DYNAMIC. Genotype is not disclosed to the participant. Study staff with direct participant contact remain blinded to genotype. Because *APOE4* does not have equal penetrance in the Clinical Cohort, the KU ADC continually monitors enrollment rates for DYNAMIC based on sex and E4 carriage and an unblinded study team member not involved in recruitment, consent, or study visit execution reviews and provides enriched contact lists to blinded staff to support balanced participation. Participants who have not previously had their E4 characterized consent to have genotyping performed for the purpose of the study. We exclude individuals with an E2 allele. We make efforts to preferentially match *APOE4* genotype groups based on sex.

## Procedures

Participants attend a single study visit. Because our focus is on assessment of dynamic time- and intervention-related changes in our outcomes, precise study timing and short transitions between study activities is critical. Our procedures have been planned to minimize waiting and transition time between study events.

Participants first change into provided magnetic resonance imaging (MRI) compatible clothes (scrubs) and remove all MRI-incompatible dental appliances, jewelry, etc. The exercise bicycle ergometer (Corival, Lode B.V., www.lode.nl) is adjusted so that the knee achieves near but not complete extension. Participants practice pedaling between 60–70 rpm until they report feeling comfortable with the movement and equipment.

Participants then complete approximately 20 minutes of NIH Toolbox-based neuropsychological testing on an iPad (Apple Inc.) in the following order, Pattern Comparison, Picture Sequence Memory, and Flanker Inhibitory Control and Attention tests. Two seated blood pressure and pulse readings and taken one minute apart are averaged as a baseline measure of vitals (Welch Allyn ProBP3400).

Next, participants are escorted to the adjacent MRI suite. They are fitted with a continuous blood pressure monitoring cuff (Caretaker 4, Caretaker Medical N.A. caretakermedical.net) on the finger which is calibrated to the baseline vitals. The MRI technologist fits ear plugs and headphones on the participant, lays them on the MRI table positioning the cuffed finger on the abdomen and begins scanning. Rapid transition from preparation to scanning is emphasized.

Imaging data are collected with a 3 Tesla whole-body scanner (Siemens Skyra, Erlangen, Germany) fitted with a 20-channel head and neck receiver coil. The MRI session is split into two parts: pre-exercise, and post-exercise. Each portion of the session begins with automated scout image acquisition and shimming procedures to optimize field homogeneity. The pre-exercise portion consists of two 3D turbo gradient spin echo (TGSE) pseudo-continuous arterial spin labeling (pCASL) sequences,(70–73) yielding 9 minutes and 56 seconds of pre-exercise pCASL data. All pCASL sequences are collected with the same acquisition parameters, designed for full brain coverage, and optimized for cerebral blood flow (CBF) measurements in older adults (repetition time/echo time (TR/TE) = 4300/22.42 ms, flip angle = 120 deg, field of view (FOV) = 300 × 300 mm^2^, matrix = 96 × 96 voxels, voxel in-plane resolution = 3.125 × 3.125 mm^2^, slice thickness = 2.5 mm, 48 axial slices, 20 volumes, acquisition time = 5:48). Positioning of the pCASL sequences is prescribed using the automated scout image, aligning the top of the acquisition box to the top of the brain and the angle of acquisition to the base of the corpus callosum as landmarks. The two pre-exercise pCASL sequences are followed by a T_1_-weighted, 3D magnetization prepared rapid gradient echo (MPRAGE) structural scan(74) (TR/TE = 2300/2.95 ms, inversion time (TI) = 900 ms, flip angle = 9 deg, FOV = 253 × 270 mm^2^, matrix = 240 × 256 voxels, voxel in-plane resolution = 1.05 × 1.05 mm^2^, slice thickness = 1.2 mm, 176 sagittal slices, in-plane acceleration factor = 2, acquisition time = 5:09), used for alignment and normalization of the pCASL data.

Participants return to the testing room. An optical heart rate sensor (OH1 Polar Electro, Inc. polar.com) is secured to the forearm with self-adhering wrap. Blood pressure is taken again. Then, a flexible intravenous catheter is placed and 10mL of blood is collected in vacutainer tubes containing EDTA as an anti-coagulant. If the genotype is not available, an additional 3mL of blood is collected in a single vacutainer tube containing acid citrate dextrose and stored for future genotyping.

Participants then remount the cycle ergometer and begin a 5-minute warm up. During the initial 5 minutes, study staff gradually increase resistance with a goal of achieving the target heart rate of 45–55% of HR reserve in minutes 4 and 5. Participants begin pedaling at 60–70 rpm and a resistance based on an age-dependent decision algorithm (Figure 1). The 15-minute aerobic exercise bout begins immediately following the warm-up. Study staff check heart rate every 1-minute and adjust cycle resistance to maintain the heart rate in the target zone. After 15 minutes of exercise resistance is reduced to 10W and participants pedal at a self-selected cadence for 3 minutes to cool down and drink 100mL of water to reduce potential perspiration-related changes in blood volume. Post-exercise vitals are taken immediately at the beginning of the cool down period. An additional 10mL of blood is collected during cool down. The heart rate monitor is removed, and the participant is quickly escorted back to the MRI room.

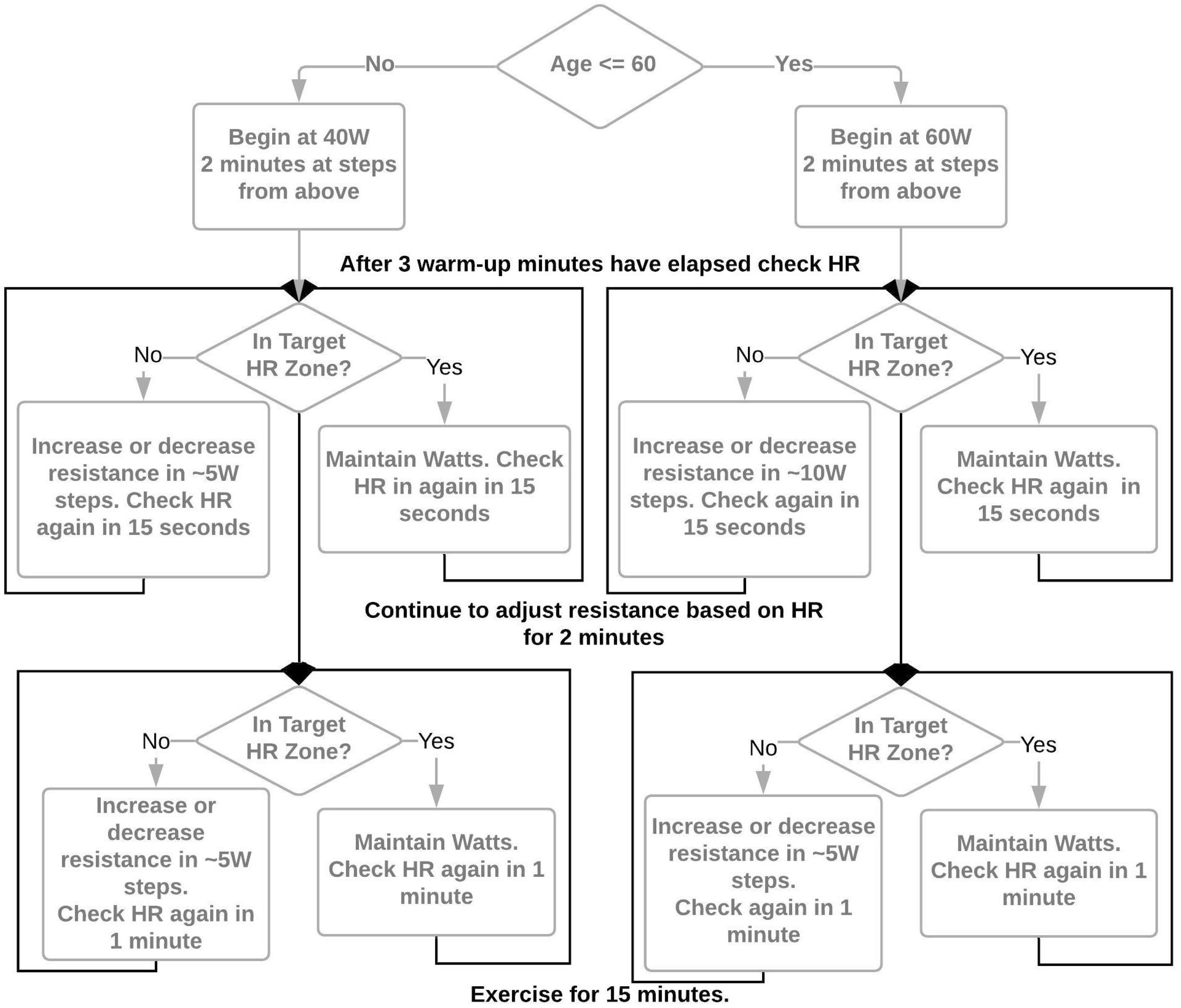

Once back in the MRI room, the same preparatory procedures for MRI are repeated and 4 consecutive pCASL sequences are acquired, yielding 23 minutes and 12 seconds of post-exercise pCASL data. Finally, the participant is escorted back to the testing room where vitals and 10mL of blood are taken one more time, and neuropsychological testing is repeated. Participants are compensated $100 upon completion of the visit.

### Blood Processing and Assessment

We optimized sample collection and processing procedures for accurate measurement of plasma neurotrophins in 5 samples independent of this study. As demonstrated in our proof-of-concept results, when platelets remain in a blood sample, a freeze thaw can greatly increase the concentration of such factors and may not accurately reflect levels that were circulating in the plasma at the time of acute exercise. Consequently, our protocol emphasizes immediate processing. Our optimized protocol is as follows: plasma is generated immediately upon collection by centrifugation by processing at 1500 RCF (g) (2800 RPM) at 4°C for ten minutes. Platelet-rich plasma is then centrifuged in four, 1.5mL aliquots at 1700 × g (4500 RPM) (4C) for 15 minutes. The resulting platelet-poor plasma is separated from the pellet and snap frozen in liquid nitrogen until stored at –80 C at the end of the visit.

### Imaging Analysis

Planned pCASL data analyses include using the USC Laboratory of Functional MRI Technology CBF Preprocess and Quantify packages,(75, 76) and Statistical Parametric Mapping CAT12 package.(77) We motion correct labeled and control pCASL images separately, calculate the CBF timeseries via simple subtraction, and coregister with the T_1-_weighted anatomical image. Mean CBF in the entre gray matter mask is extracted from each label-control subtraction image in the timeseries in units of mL*100g tissue^−1^*min^−1^. Imaging analysis may change as the field improves methods.

### Cognitive Test Assessment

Cognitive assessments scores are calculated automatically by the NIH Toolbox software https://www.healthmeasures.net/). We plan to use change in T-Score for each domain which is age, education, gender, and ethno-racial identification corrected and provides a score based on a normative mean of 50 with a standard deviation of 10.

### Data Collection

All data are collected and organized in a custom designed REDCap(78) database. Project access is role based. APOE4 genotypes are kept in a separate database and the linking list is kept by a designated, unblinded investigator.

### Sample Size

To our knowledge, there are currently no peer-reviewed reports of genotype-based CBF differences in response to acute exercise. However, perfusion measures in genetic risk for AD (*APOE4*) have been performed previously and can form the basis of a reasonable power analysis. Two prior cross-sectional estimates of the relationship of perfusion and E4 carriage have delivered similar effect sizes *(d* = 1.0).(48, 49) Given this effect size, we expect to be able to discern differences based on *APOE4* in a sample of 60 older adults.

### Safety

#### Adverse Events

Adverse events are defined as any untoward medical occurrence in study participants, which does not necessarily have a causal relationship with the study treatment. The seriousness of the adverse event is determined using the National Cancer Institute Common Terminology Criteria for Adverse Events (CTCAE) v3.0, and need for hospital admission. Adverse events are assessed only at the visit, but the consent form has contact information should the participant need to contact the study team regarding delayed development of an AE. Serious Adverse Events are reported per institutional and NIH requirements.

#### Monitoring

Safety of the study is monitored in an ongoing manner by a chartered Data and Safety Monitoring Committee (DSMC) and Independent Safety Officer according to a Safety Plan. The Independent Safety Officer advises the NIH and the Principal Investigator regarding participant safety, participant risks and benefits, scientific integrity and ethical conduct of a study. The DSMC provides additional support and guidance for the Principal Investigator.

### Response to SARS-CoV-2

Data collection began prior to the SARS-CoV-2 novel coronavirus pandemic, was paused between March 11 and June 1, 2020, and resumed with additional safety procedures. The blood processing centrifuge and staff member were moved to a separate, nearby testing room to allow for physical distancing. All participants are screened via telephone a maximum of 72 hours before the day of visit, and are screened again (including temperature) outside the imaging facility on the day of the visit. All staff wear level 1 surgical masks, gloves, and face shields. Participants are provided a surgical mask to be worn at all times except during exercise and in the MRI. The staff member in charge of exercise and neuropsychological testing minimizes time within 6 feet of the participant and MRI tech staff. Electronic, advance consenting has been implemented following the coronavirus pandemic to reduce the amount of close contact time between staff and participant.

## Proof-of-Concept Results

To date, we have enrolled 16 participants into the study. Demographics are provided in Table 1. To demonstrate proof-of-concept, we provide preliminary analyses and comparisons of individuals above and below 65 years of age. E4 carriage has not been unmasked to date and is not included in this report. There were more women in the older adult group. Figure 2 depicts the study flow and average time for each study event. Figure 3 demonstrates our ability to capture dynamic blood flow changes post-exercise. The area under the CBF curve (Figure 3A) is greater in the younger individuals. Additionally, even when we normalize CBF to the average of the second pre-exercise pCASL sequence (a stabilized resting CBF), the area under the curve of younger adults remains greater. Notably, these dynamic changes do not appear to be heavily driven by blood pressure which is plotted underneath the CBF curves.

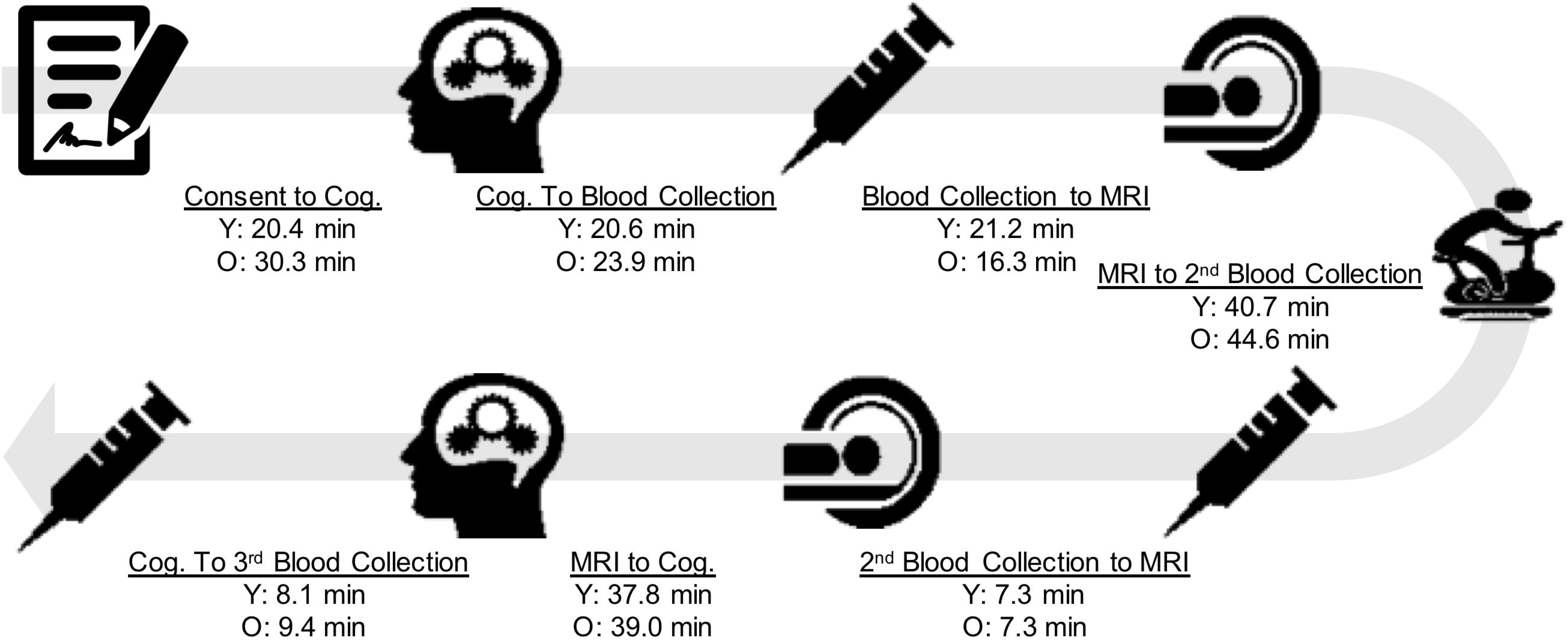

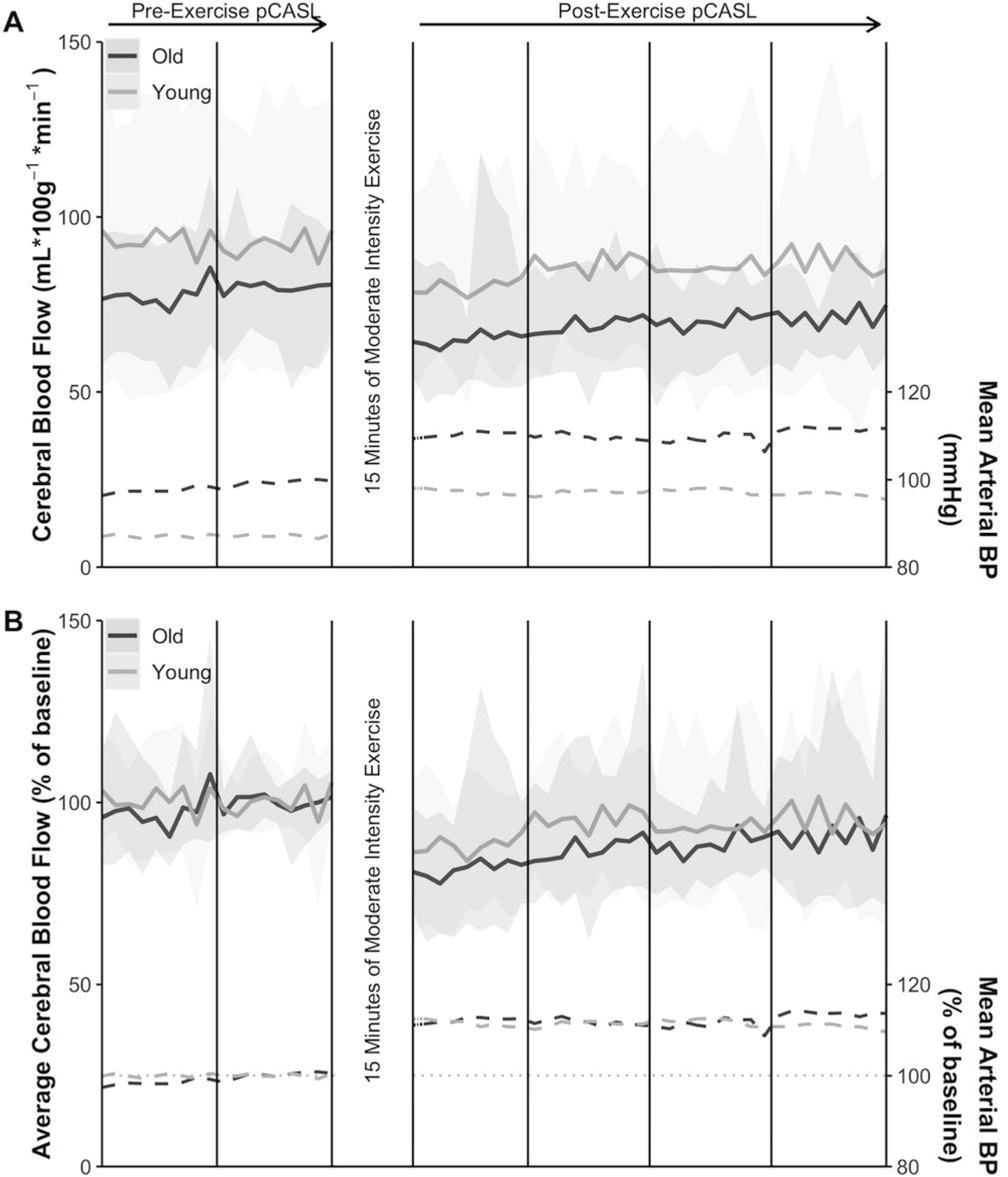

We also provide evidence for our decision to process blood collection immediately. In plasma processed with one centrifugation, mean plasma levels of BDNF were 17,506 ± 4031 pg/mL. Duplicate samples that underwent an immediate second centrifugation to remove the platelet pellet, followed by an immediate snap-freeze, were measured as having mean levels to 29 ±10.6 pg/mL. A delay of 15 minutes prior to each centrifugation step, which may allow increased time for platelet release when activated by the shear stress of centrifugation, increased levels to 245 ±108 pg/mL.

### Adverse Events

To date there has been one adverse event, nausea, upon IV placement. Symptoms resolved with a light snack and rest.

## Discussion

We have designed and implemented a single visit clinical trial to test the effect of *APOE4* carriage on brain blood flow response to an acute, 15-minute, moderate intensity exercise challenge. We have also refined optimal blood collection and processing procedures to characterize circulating, bio-available neurotrophic responses to exercise. We expect that our strict and time sensitive protocol will allow us to identify CBF and blood-based neurotrophic responses that are obscured during typical resting conditions. We will also explore whether CBF or neurotrophic responses are related to performance changes on neuropsychological tests. To be clear, we do not expect measurable vascularization, neurogenesis, or other benefits immediately following exercise. Nor do we suggest that any neurotrophic increases are causal of *ad hoc* cognitive change or CBF response. Rather, we seek to index the transient changes and relationships that are hypothesized to mediate these benefits with chronic exposure to exercise.

Our proof-of-concept results suggest that we can capture a dynamic CBF recovery response following exercise that can differentiate groups. We found that older adults have lower overall blood flow and slower return to resting blood flow as measured by area under the CBF curve. Our blood processing results provide a clear case for immediate post-processing to identify the bio-available circulating neurotrophic factors following exercise. Optimization of preand post-processing techniques revealed higher BDNF levels in plasma collected without removal of platelets compared to plasma where platelets had been removed. In addition, sitting time after the centrifugation step to remove platelets, which may result in platelet activation, also affected neurotrophic factor levels.

Despite the many strengths and carefully constructed protocol, there are notable limitations. First, our pre-post exercise measures are proxies of CBF during exercise as we are not imaging during exercise. Some protocols for MRI during exercise are beginning to emerge, though concerns about motion artifact remain. CBF can also be measured using contrast-enhanced MRI, TCD, or positron emission tomography (PET). TCD has the advantages of temporal resolution and ease of use during exercise, but can only index blood velocity. Xenon PET imaging can produce whole brain CBF but requires a radioactive isotope. pCASL provides both the whole brain spatial resolution, and potentially improved temporal resolution compared to PET, using only magnetically labeled arterial blood water, an endogenous tracer that is highly reproducible.(70) Additionally, optimal imaging parameters to capture CBF response to acute exercise using have been investigated previously.(79) Pseudo-continuous ASL sequences have improved signal-to-noise ratio while maintaining high labeling efficiency.(80) Common to all ASL sequences are pairs of images with and without labeling, that allow for CBF quantification. Typically, the subtraction of these pairs is averaged over a sequence to calculate CBF. However, the pairs viewed as a timeseries, may also capture information about transient CBF changes, for example following exercise, as we have done here. We believe this timecourse will provide important additional information about dynamic responsiveness, or cerebrovascular reserve, of the system.

A second limitation is our focus on three neurotrophic factors and the *APOE4* genotype. Recent evidence suggests that there are myriad important biochemical changes in response to exercise.(81) Similarly, thousands of genes are responsible for exercise response.(82) We have chosen to focus on those that are most consistently linked to brain health and Alzheimer’s disease in the literature. While important for our own research line, it leaves the potential for uninvestigated responses that may be important.

Despite these limitations they DYNAMIC study offers a blueprint for a unique and innovative methodology to capture acute exercise response in individuals at risk for Alzheimer’s disease. Aerobic exercise is among the most important and cost effective tools available for chronic disease management.(83) Our work along with others’ strongly suggests that moderate-intensity aerobic exercise benefits brain health and cognition.(10, 11, 84–87) However, the field continue to struggle with adequate methods for capturing mechanistic drivers of exercise benefits on the brain. New protocols such as DYNAMIC are necessary to move forward our mechanistic explorations of exercise effects on brain health and cognition.

**Table 2:** Demographics and Preliminary Results

| Table 2. Demographics and Preliminary Results |  |  |
| --- | --- | --- |
|  | Younger (n=9) | Older (n=7) |
| Age, yrs | 25.9 [23-29] | 76.6 [71-81] |
| Female, % | 5 (56%) | 6 (86%) |
| Resting Heart Rate, bpm | 81.4 [68-95] | 62 [54-71] |
| Resting MAP, mmHg | 99.6 [83.8-128.0] | 113 [90.2-142.0] |
| Mean Workload | 82.3W [49.3-114] | 29.7W [0-49.3] |
| Peak Workload | 89.3W [60-120] | 31.4 [0-50] |
| Pre-exercise MAP AUC | 1480 [1476-1485] | 1673 [1443-1913] |
| Pre-exercise MAP AUC | 3397 [3192-3602] | 3856 [3514-4207] |
| Pre-exercise CBF AUC | 1571 [920-2241] | 1339 [1043-1591] |
| Post-exercise CBF AUC | 2973 [1893-4027] | 2413 [2035-2897] |
| Values mean [range] unless otherwise noted. bpm= beats per minute;<br>CBF=cerebral blood flow; MAP = mean arterial pressure; AUC=area under curve. |  |  |

## Data Availability

Data may be obtained by contacting the corresponding author. Data will not be released prior to the closing of the trial.

## Abbreviations

AD: Alzheimer’s disease
APOE4: apolipoprotein e4
CBF: cerebral blood flow
TCD: transcranial Doppler
BDNF: Brain Derived Neurotrophic Factor
VEGF: Vascular Endothelia Growth Factor
IGF1: Insulin-like Growth Factor 1
AUC: area under the curve
KU ADC: University of Kansas Alzheimer’s Disease Center
MRI: magnetic resonance imaging
TGSE: turbo gradient spin echo
pCASL: pseudo-continuous arterial spin labeling
MPRAGE: magnetization prepared rapid gradient echo
CTCAE: Common Terminology Criteria for Adverse Events
DSMC: Data and Safety Monitoring Committee
PET: positron emission tomography

## Declarations

### Ethics approval and consent to participate

All participants have provided written informed consent approved by the University of Kansas Institutional Review Board, consistent with the Declaration of Helsinki.

### Consent for publication

Not applicable.

### Authors’ contributions

EDV, JKM, PL, SAB, JMB, LM and RJL, contributed to the conception of the work. All authors contributed to the design of the work. DW, CSJ, AK, BT, RJL, EDV, JKM contributed to the acquisition or analysis of the work. CSJ, RJL, SAB, JMB, JKM, EDV contributed to the interpretation of data. All authors have contributed to the drafting and revision of the work. All authors have approved submission and agreed to be personally accountable for the work

